# Visualising Emergency Department Wait Times; Rapid Iterative Testing to Determine Patient Preferences for Displays

**DOI:** 10.1101/2022.03.30.22273211

**Authors:** Katie Walker, Eden Potter, Indae Hwang, Tim Dwyer, Diana Egerton-Warburton, Keith Joe, Jennie Hutton, Sam Freeman, Daphne Flynn

**Affiliations:** Casey Hospital, Monash Health; School of Clinical Sciences, Monash University; Monash Design Health Collab, Monash University; Faculty of IT, Monash University; School of Clinical Sciences at Monash Health, Monash University; Emergency Department, Cabrini; MADA, MIME, Monash University; St Vincent's Hospital, Melbourne; University of Melbourne; St Vincent's Hospital, Melbourne; Sensilab, Monash University

**Keywords:** Emergency services, hospital, waiting lists, vulnerable populations, patient-centred care, decision making, information visualisation, data displays, infographics, human centred design, temporal uncertainty

## Abstract

Visualising patient wait times in emergency departments for patients and families is increasingly common, following the development of prediction models using routinely collected patient demographic, urgency and flow data. Consumers of an emergency department wait time display will have culturally and linguistically diverse backgrounds, are more likely to be from under-served populations and will have varied data literacy skills. The wait times are uncertain, the information is presented when people are emotionally and physically challenged, and the predictions may inform high stakes decisions. In such a stressful environment, simplicity is crucial and the visual language must cater to the diverse audience. When wait times are conveyed well, patient experience improves. Designers must ensure the visualisation is patient-centred and that data are consistently and correctly interpreted. In this article, we present the results of a design study at three hospitals in Melbourne, Australia, undertaken in 2021. We used rapid iterative testing and evaluation methodology, with patients and families from diverse backgrounds as participants, to develop and validate a wait time display. We present the design process and the results of this project. Patients, families and staff were eligible to participate if they were awaiting care in the emergency department, or worked in patient reception and waiting areas. The patient-centred approach taken in our design process varies greatly from past work led by hospital administrations, and the resulting visualisations are very distinct. Most currently displayed wait time visualisations could be adapted to better meet end-user needs. Also of note, we found that techniques developed by visualisation researchers for conveying temporal uncertainty tended to overwhelm the diverse audience rather than inform. There is a need to balance precise and comprehensive information presentation against the strong need for simplicity in such a stressful environment.

## 1 Introduction

Emergency departments (EDs) treat a large volume of patients daily. In Australia, EDs provide approximately 31,000 visits per annum per 100,000 people, in the US there are about 43,000 visits per 100,000 [4,39]. Patients arrive hoping to see a clinician immediately, but usually have to wait for care. Approximate wait times are important to patients and families [41]. Before attending, online wait time displays can be used to inform a decision about which facility to attend [36]. Once in an ED, wait estimates are used to meet patient emotional, logistic and physical needs and also assist paramedics whilst patients wait on stretchers [41]. Wait times can be estimated using prediction models, with varied accuracy [2, 37, 40, 42]. Despite the high volume of patients exposed to ED waits, when displays are designed there is little to no input from patients and families who are the end users of the displays. Displayed information about wait times is important as high stakes decisions may be made by patients in response, such as leaving before being attended by a physician or rearranging multiple external commitments [41]. Additionally, patients are having to make these decisions in what is an emotionally and physically stressful environment. During the COVID-19 pandemic, wait times have been long, and patients have had to decide whether to attend an ED at all; if they attend, whether they should risk infection by waiting for hours in a waiting room alongside people who may have COVID-19; and whether there is an alternative ED they can attend with a shorter wait time.

There are a range of ED wait time displays that have recently been made available online to consumers, Fig. 2. Some are designed for health administrators or report system wide performance, others are targeted at consumers, with various amounts of information visualised in different ways. Some wait time displays are comparative, often displayed in a fee-for-service ED operating environment, others display information only applicable to a single ED. To our knowledge, there is no patient-centred design research available to guide health administrators when they consider displays for their institutions. In Section 2 we review past research on data visualisation, infographic design, cognitive load, and visual perception, but argue that this past work leaves a gap when it comes to the considerations for presentation of wait time information. Even if the wait time presented to consumers is accurate, current displays may or may not be useful to consumers and may or may not be understood by patients and families from diverse demographics with varied data literacy levels.

**Fig. 1.**
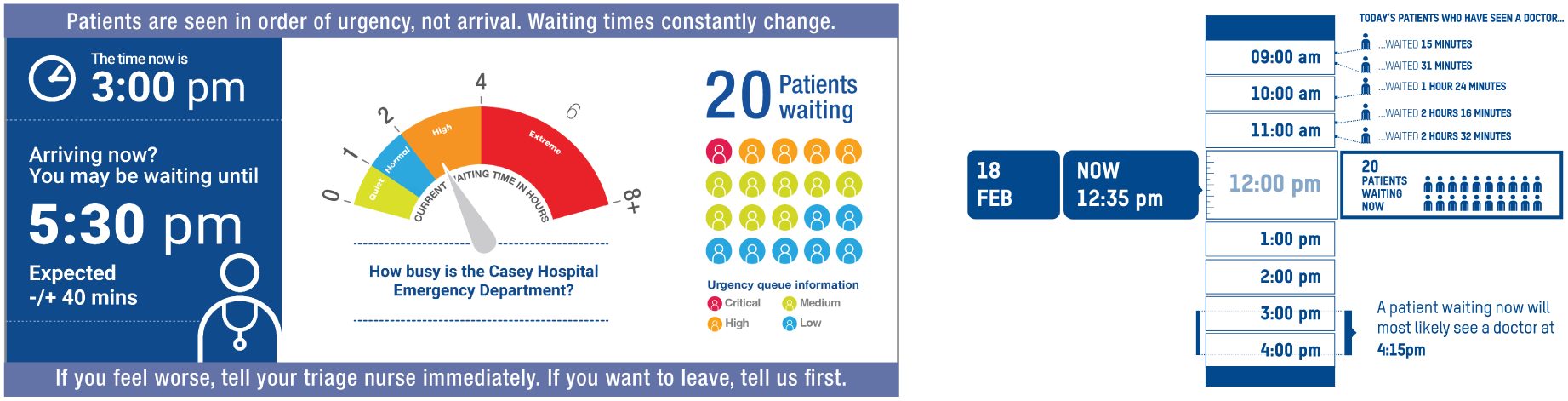
Following a Rapid Iterative Testing and Evaluation (RITE) methodology, we obtained qualitative feedback from actual waiting patients on a variety of visualisation elements and styles in our study. These visualisations ranged from relatively straightforward displays relying on textual information (left) to displays replacing text with visual explanation as much as possible, for example, using a spatial representation of time and the arrival and wait time information upon which the estimated wait time window is based (right).

**Fig. 2.**
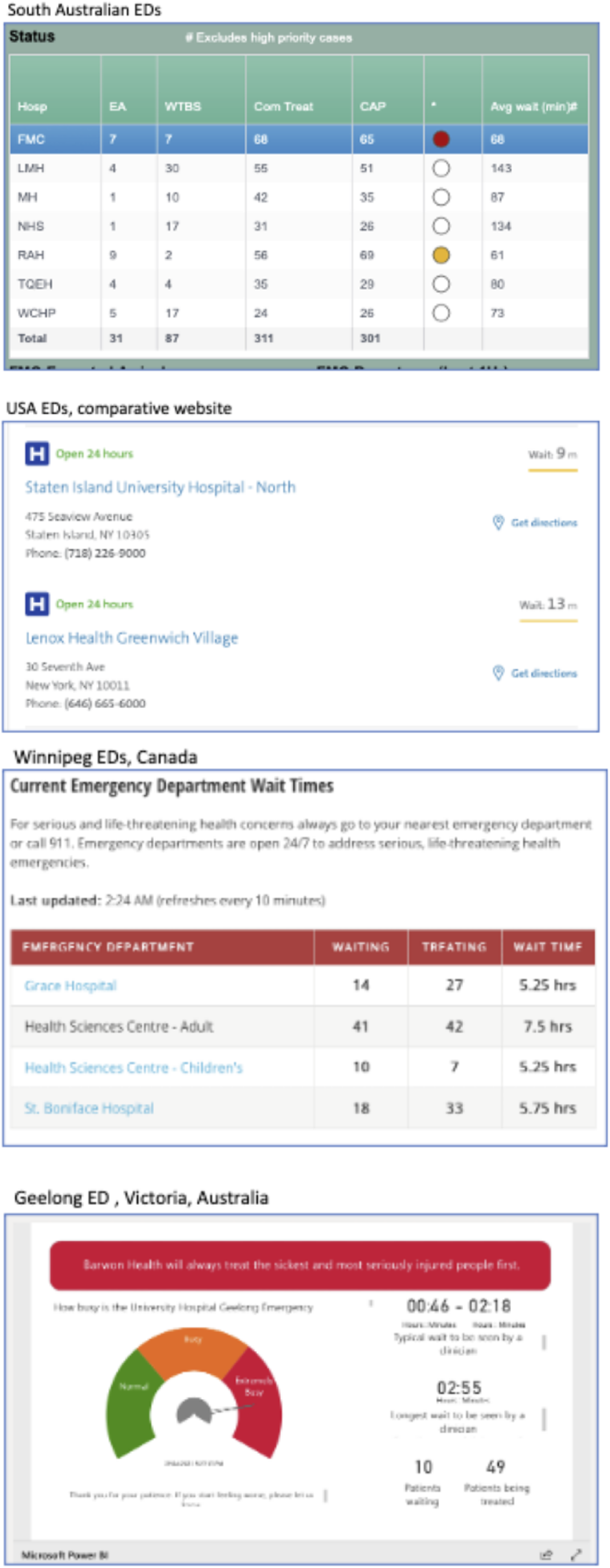
Comparison of currently publicly available online ED wait time displays. Information presented is minimal and generally presented textually rather than visually.

As described in Section 3, we deployed human-centred design to evaluate how ED wait time displays could be useful and understandable to patients and other visitors to the ED. Human-centred design is a mindset and approach that foregrounds the needs and experiences of the people who will use or interact with the designed service, system or product. This stands in contrast to designer-centric approaches, where the designer’s creative intent drives both the interpretation of the design problem and its resolution through design [13]. The techniques and activities used in the human-centred design process are focused on understanding the intended audience or user group, and designing specifically for this.

Our study aimed to visualise ED wait times in a display that is useful to patients and families and that can be understood by a range of people from culturally, linguistically and educationally diverse backgrounds.

## 2 Background

From a visualisation perspective in the era of “big data”, displaying a few small pieces of temporal and quantitative information (e.g. number of patients currently waiting, and expected wait time), may seem like a straightforward requirement for a visual design. Visualisation research typically focuses on developing sophisticated display and interaction techniques suitable for large and complex data for highly trained users, such as data scientists or domain experts. More recently, “personal visualisation” has focused on a more general user-base, aiming to deliver data relevant to individuals’ own lives through their personal electronic devices [32]. While a broad variety of data has been considered in this context, ranging from useful and timely (e.g. bus wait times [25], general health and fitness) to whimsical and artistic (e.g. Dear Data) [27], we note that most work in this area (see [1] for full survey) considers a relatively sophisticated user-base, i.e. people willing to use their electronic devices to assist with relatively complex tasks.

As a use-case of data visualisation, designing public information displays for an ED represents a significantly different design challenge compared with those typically considered in the visualisation literature. The data is minimal, as already discussed, but has hidden subtlety, as we discuss shortly. The potential users of the displays are extremely diverse and include the elderly; people with minimal literacy or intellectual disabilities; people who are intoxicated or experiencing mental health issues; and people for whom the English language is a challenge. Research in data visualisation targeting such a diverse user base is currently lacking, but we would argue that research targeting a broader user base (e.g. not just targeting data scientists, domain experts, or others with university-level education) could make data visualisation accessible to billions more users around the world [23].

In an ED wait room, many (if not most) users of displays are likely to be distressed. It is highly important that the information presented is properly understood. If patients misunderstand the information, or are overwhelmed by it, they may make a poor decision, such as leaving before they are assessed by a health professional or engage in other harmful behaviour. In other words, this is a high and immediate consequence application of visualisation. In Fig. 3, we visualise the potential target domains of data visualisation on three axes, broadly characterising the data complexity, potential user base and consequentiality of the application. We see that ED wait time displays are an outlier across all three dimensions.

**Fig. 3.**
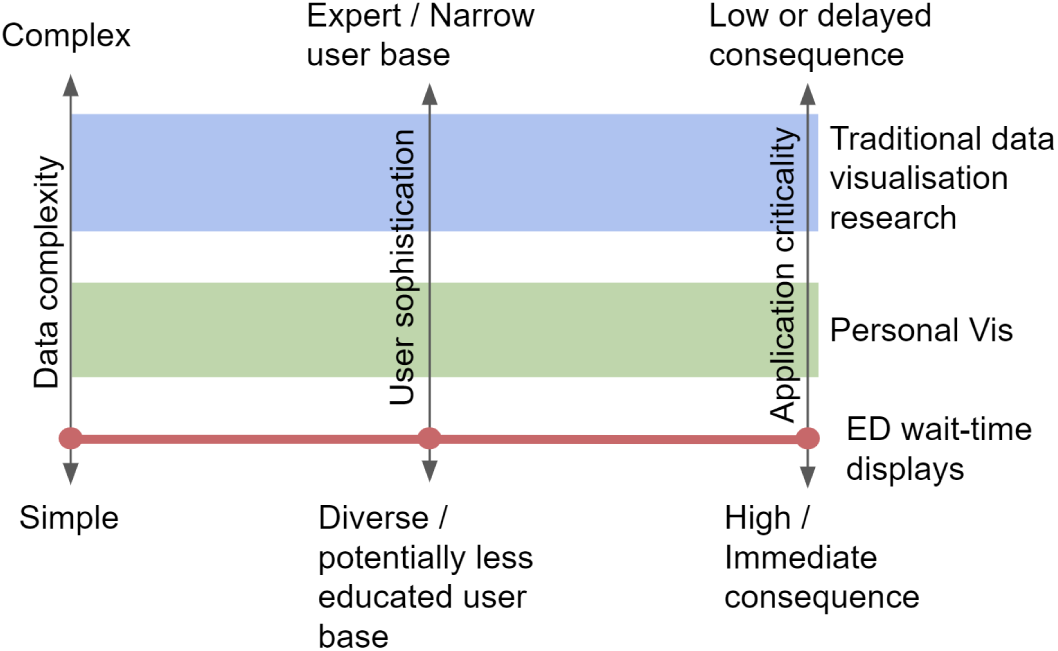
The space of target domains for data visualisation broadly characterised by three dimensions of data complexity, user sophistication and application criticality. ED wait time displays are an outlier across all three dimensions as indicated by the red line.

Rjabiyazdi et al. characterised communicating health data to patients as a “wicked problem” [34]. However, their focus was designing personalised data displays that could be mediated by an expert agent (i.e. a health professional) to cater to the patients’ individual condition, and their own ability to engage with complex information. The ED wait context, while carrying similar criticality in terms of needing to guide patient decisions with significant health outcomes, poses additional challenges. While it is conceivable that the information could be tailored to individuals via their personal device, in practice the displays need to be available to people without access to devices or without the capability to use an electronic device.

This highlights the importance of the concept of “Techquity” [35]. That is, “the strategic development and deployment of technology to advance health equity”. In an era of COVID-19, the increased burden of illness has fallen disproportionately on those with the least resources. At such a time, the priorities of health technology should be processes and outcomes that address health equity and social justice. In the context of the emergency waiting room the most disadvantaged patients are more likely to leave before being seen, and it is these patients that could most benefit from good waiting room design and information.

Through the course of our design and evaluation process, the following issues and research questions emerged that we believe make it an unusual use-case which standard data visualisation design guidelines (e.g. [31]) do not completely inform.

As described in Section 3.7, the precise set of information to be displayed is open. There is much information available from the patient check-in system that could be displayed – but some information could be harmful (triage status of individuals, etc), and too much information could overwhelm. Thus, our user-centred design question was not just “how” to show the data, but also “what” to show for the best interests of both patients and staff.

The wait time is an estimate from an AI model and comes with an estimate of accuracy (e.g. a most-likely time window). Conveying uncertainty through visualisation is another open challenge - even in expert domains [11]. We want to build “trust” but we do not want to build an expectation which may not be met. There is past work indicating that the form of a visualisation can influence decision making under uncertainty [5], but this was an artificial, low-stakes scenario involving young mostly educated technologically adept people (i.e. Mechanical Turk users) []. Does such best practice for uncertainty visualisation, particularly temporal uncertainty, translate into the high-stakes environment of an emergency department or should we follow a different norm of simply leaving the uncertainty implicit [19]?

## 3 Methods

### 3.1 Study design

Emergency stakeholders partnered with design and visualisation experts to determine elements of wait time displays that might be important. The full timeline charting our activity is summarised in Fig. 4. Designs were developed that might convey these elements to consumers. Rapid Iterative Testing and Evaluation (RITE) [28] methodology was used between March and August 2021, inviting emergency patients, accompanying family members and patient reception staff to review paper versions of displays and provide interpretations and feedback on each element. Participant feedback was applied to features of the design in the form of “fixes” [28], and the new iteration was again shown to participants working in or visiting the ED to gain further insights until a consensus and “best version” of the display was obtained. Ethics approval was obtained from Cabrini Human Research Ethics Committee (10-13-05-19).

**Fig. 4.**
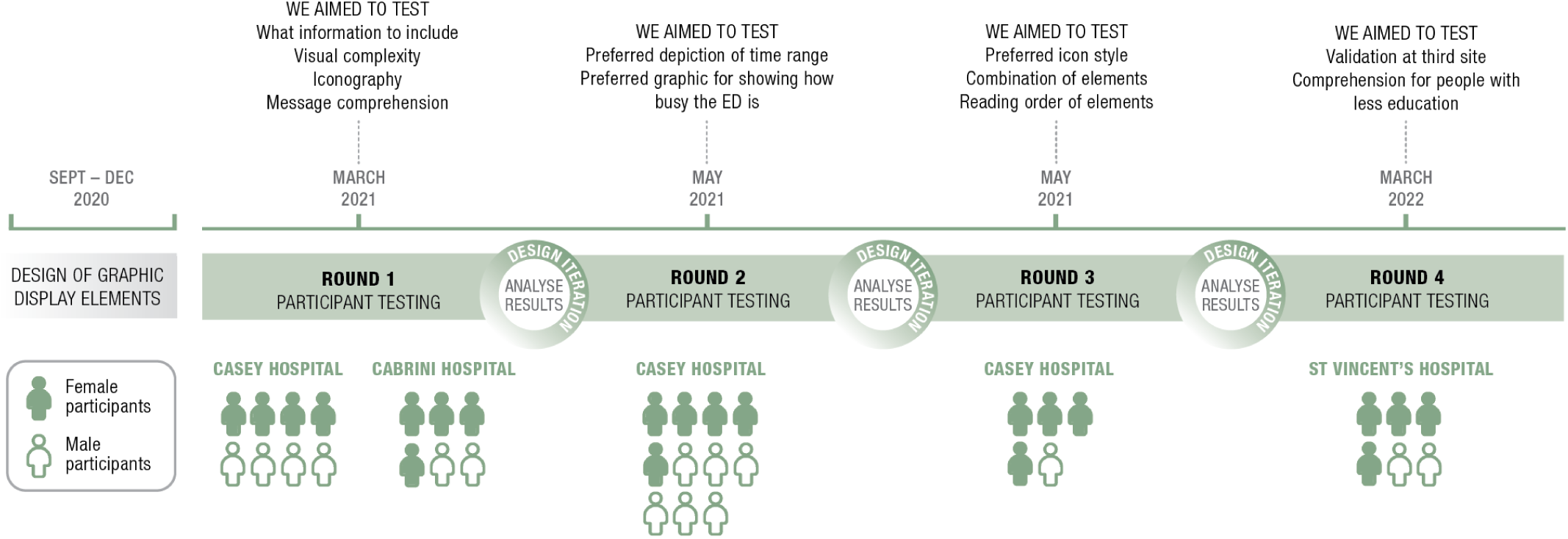
Timeline of activities and testing iterations from September 2020 to March 2022

### 3.2 Initial designs

Emergency Department wait time displays from around the world, currently available on the internet, were reviewed regarding variables and types of displays. The design team included emergency physicians, interaction designers, health design, and human-centred computing academics, who worked together to determine the take-home message for consumers intended by the visualisation and possible variables available to be used. Potential individual components of the visualisation were determined and designs to convey each datapoint were developed by an academic design artist (IH). A series of meetings were held to refine designs until a consensus was achieved amongst the design group regarding components and designs to be tested. Iterative improvements were made to designs and new sets of images and questions were developed, checked by the team and tested following each meeting. This process continued until the team felt that enough iterations had been undergone to recommend the displays to EDs.

### 3.3 Rapid Iterative Testing and Evaluation (RITE) methodology

The resources for this study were limited, requiring the time contributions of extremely busy ED staff and access to patients was made more complex, as it was conducted during a time of heightened COVID-19 restrictions. We therefore needed a study methodology that would allow us to get feedback for as many design ideas as possible and quickly eliminate the designs that did not show promise.

Rapid Iterative Testing and Evaluation methodology is a way of rapidly refining and improving designs or software, based on direct consumer or user feedback [29,45]. Users are asked to review or use designs and comment on them at the same time. Modifications to designs are made quickly, sometimes after only one participant’s feedback and then new iterations are tested. There is rapid cycling between design and testing based on user feedback, allowing participants to provide input at each stage of software development. This process continues until most or all solvable issues are resolved.

This study used the RITE methodology to rapidly test and modify wait time visualisations, discarding the least preferred or confusing elements, whilst retaining the elements considered to be the most useful, clearest and important. After individual elements were tested and finalised, participants were invited to review combined displays until finalised. If there was equipoise amongst participants regarding an element, either option would be recommended for live visualisation.

### 3.4 Settings and participants

Three EDs in Victoria, Australia contributed data. The EDs were at Cabrini, Casey and St Vincent’s hospital. These public and private EDs serve a wide range of patients. Two treat a culturally and linguistically diverse (CALD) younger population with lower affluence compared to the rest of Victoria. The other caters for mainly English speaking, older patients with high levels of private health insurance and health literacy. Levels of data literacy and comfort with technology vary widely across their catchments. Casey and St Vincent’s EDs receive block, per-annum, governmental funding (no per-patient payments). These hospitals have no incentive to advertise to attract more patients and often experience high levels of ED crowding. At the time of the study, they did not display wait times. Cabrini ED is a private, fee-for-service facility and had a wait time estimate displayed in the waiting room and online during the study (Fig. 5).

**Fig. 5.**
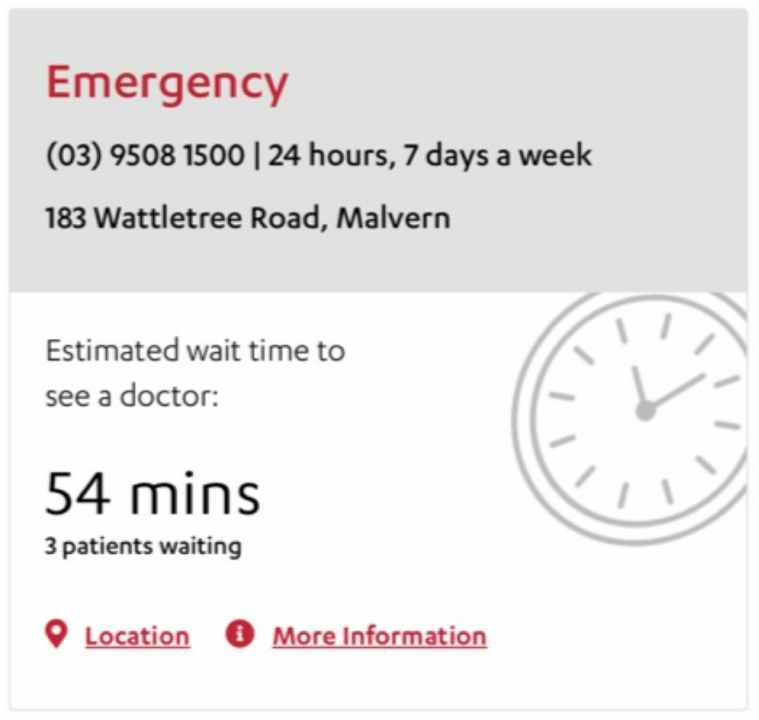
Cabrini wait time display during research period

### 3.5 Eligibility

Patients and accompanying support persons were recruited from ED waiting rooms, treatment areas and short stay units after triage had been completed. Triage nurses and reception clerks were recruited from acute ED areas. People were eligible for inclusion if they were waiting for care, results or inpatient bed allocation, as phone interviews could not delay clinical care. They had to be clinically stable, calm, deemed at low risk of COVID-19 and able to provide informed consent. People had to be capable of looking at paper images and speaking on their telephone. Interpreters were not available, so to incorporate the views of non-English speaking patients and families, family groups could be interviewed, with an English speaker eliciting the views of the nonEnglish speaking family members and translating their responses. The recruitment of family groups also allowed for people with other barriers to participation to be included (such as difficulties with communication, telephones or turning pages).

### 3.6 Intended users and content delivery methods

The displays are intended to be consumed primarily by patients and their support persons and/or family members. They will also be used by arrivals staff (triage nurses, paramedics) and clerks. Content was intended to be streamed initially to screens in waiting rooms, then the internet and finally texted to patients’ mobile phones at check-in if no issues were raised with the first delivery methods.

### 3.7 Variables available for display

Prediction models were derived from variables available via the Victorian Emergency Minimum Dataset (VEMD) [9]. This dataset is required of all EDs in Victoria and includes demographics and patient flow variables. Some of these variables are available after patient registration and triage. Calculated variables can be generated based on these data relating to patient flow, such as the number of patients in the waiting room, being treated and awaiting an inpatient bed. The model output predicts the time from triage-to-clinician. The accuracy of the model is approximately +/-20-40 minutes depending on the complexity of the ED and whether an ED uses a global or customised prediction model [40].

### 3.8 Recruitment and participation

Purposive sampling was undertaken until participants from a broad spectrum of demographics had participated in the study. Characteristics sought included males and females, a range of ages, occupations, ethnicity and languages spoken at home. Varied levels of education, data literacy, familiarity with EDs, disability/ability, and participant roles related to the ED visit (patient, parent, child, spouse, friend, other caregiver) were also sought.

A researcher identified patients who might be suitable to participate on a convenience basis, at times when the interviewer was available (KW, KJ, SF). Potential participants were approached face-to-face, the purpose of the study was explained, written patient information and consent forms (PICFs) and visuals packs (containing images for testing) were provided. Permission was sought for a researcher to call them. The physician also explained that the healthcare team would not be aware of any part of their interview and they were encouraged to speak freely about the topic (whilst avoiding any personal health information). The researcher (EP) called a few minutes later (to allow PICF reading time), explained that they were not a healthcare worker, obtained verbal consent and conducted a telephone interview. The interview could be interrupted at any time and healthcare needs were prioritised.

### 3.9 Data collection and analysis

After initial demographic questions, the interviewer directed participants to look at specific pages in their visuals pack, describe what each visual was communicating and then discuss preferences for displays based on elements shown to them on each page. Questions asked participants to rate visuals out of five (where a score of one was “not at all useful” and five was “very useful”), and were asked to suggest why they allocated a score to a given visual. Interviews were audio recorded and new interviews continued until the interviewer (EP) felt that no new data had been obtained in the preceding two interviews or that there were such great design flaws identified by participants that the images needed reworking prior to further testing. Recordings were transcribed by the interviewer. Transcripts were not returned to participants for checking. Data were extracted and analysed by one researcher (EP) using a data table [8] to organise and compare results. Findings for each round of testing were conveyed to the design team at a team meeting, and a plan was made for future testing as needed.

## 4 Results

### Participant flow and demographics

Initially, there were three groups of participants (n=14,11,5) recruited onsite in two different EDs over a 4 week period. Each group was shown a different set of wait time screen designs, with fewer design options produced and also fewer interview questions in each cycle as the iterations progressed and the team refined the design details. Key insights milestones for each iteration are shown in Fig. 4. A final round of testing (participant n=6) in a third hospital with a wide cultural demographic served to test information comprehension for the refined versions. The demographics of the participants that were recruited are reported in Table 1.

**Table 1.** Demographic breakdown of our participants (n=36; *=reflects one participant not being asked a question due to healthcare needs)

|  | Number | % |
| --- | --- | --- |
| **Gender** |  |  |
| Male | 15 | 42 |
| Female | 21 | 58 |
| **Age** |  |  |
| Teens (13-19) | 1 | 3 |
| 20-29 | 4 | 11 |
| 30-39 | 6 | 17 |
| 40-49 | 13 | 36 |
| 50-59 | 5 | 14 |
| 60-69 | 2 | 6 |
| 70-79 | 5 | 14 |
| **Language spoken at home\*** |  |  |
| English | 30 | 83 |
| Arabic | 1 | 3 |
| Assyrian | 1 | 3 |
| Greek | 1 | 3 |
| Italian | 1 | 3 |
| Hindi and Punjabi | 1 | 3 |
| **Highest level of educational attainment\*** |  |  |
| 9th to 11th grade | 5 | 14 |
| High school graduate | 5 | 14 |
| Some college, no degree | 9 | 25 |
| Bachelor's degree | 11 | 31 |
| Masters, professional or doctoral degree | 5 | 14 |

### Testing and refinement

Several sets of ED wait times screen visuals were designed for consumer testing. The main goal in designing visual elements for each ED wait time visual was to support message comprehension for patients and visitors from multicultural backgrounds and varied cognitive abilities. Our icon design was based on the public information symbol set in the International Language of ISO graphical symbols, a standardised set used to support or replace written language forms [21, 22]. Symbols and icons have been used in health-related information displays for almost 100 years, starting with Otto Neurath’s picture language system ISoTyPE, devised to communicate social and economic data of the day so that people would understand them and be more likely to remember them [33].

The visuals’ colour palette was chosen according to local and international cultural conventions around colour connotations. For example, red is used to suggest highest urgency and green to connote stability and safety in Australian fire warnings, traffic light systems, and weather maps. In addition, we modified the tone of colours to improve the distinction of the four primary colours for colour blind accessibility. Font choices considered the cognitive-perceptual aspects of readability and legibility. Sans-serif typefaces – those without decorative elements – were selected to promote maximum readability on digital display screens [18].

#### 4.0.1 Wait duration or absolute time to be seen?

The first set of designs presented to participants in Round 1 concerned how best to present wait time relative to current time. The three alternatives considered included: (A) an absolute time range to be seen; (B) predicted wait time with a +/ − time window estimate; (C) absolute time to be seen and time window estimate (below).

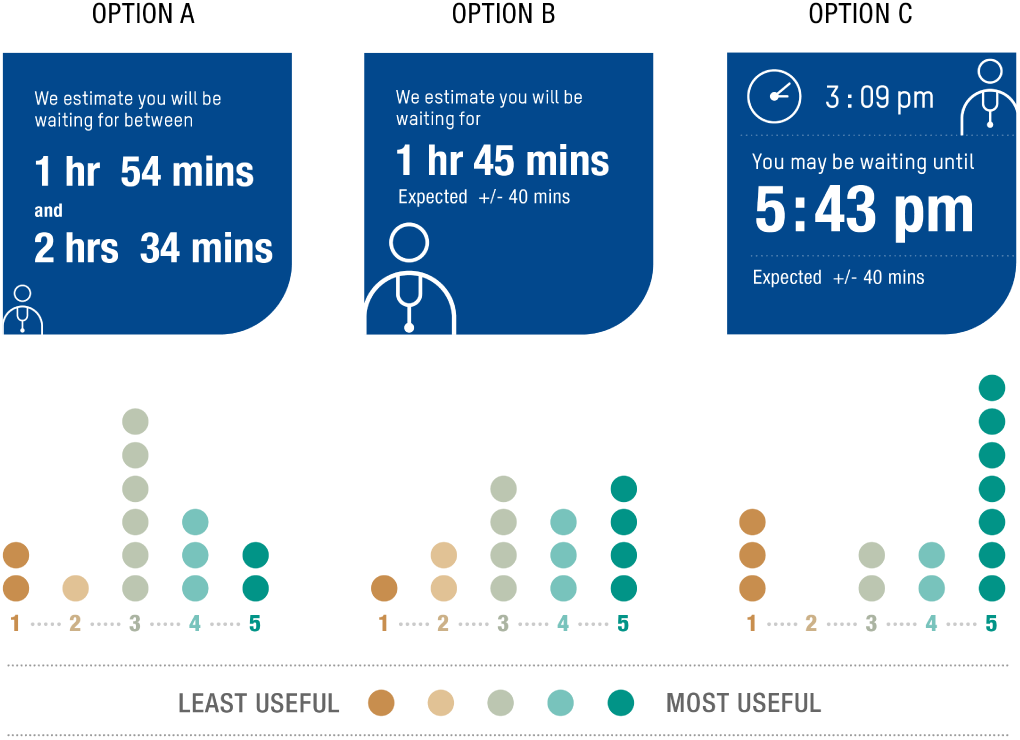

Most people, including all aged over 70 years preferred to see the current time and the expected time to be seen (Option C) rather than wait duration. They preferred to see both an analogue and a digital clock form of the time. It was “easier to orientate yourself”. Some participants struggled to work out when they would be seen if given only the wait duration. People perceived Option A, the estimated “between” duration of waits as longer waits than if the times were calculated for them and were somewhat depressed by the content.

#### 4.0.2 Images or words?

Also in Round 1, we compared options that varied the degree to which text was replaced with visuals.

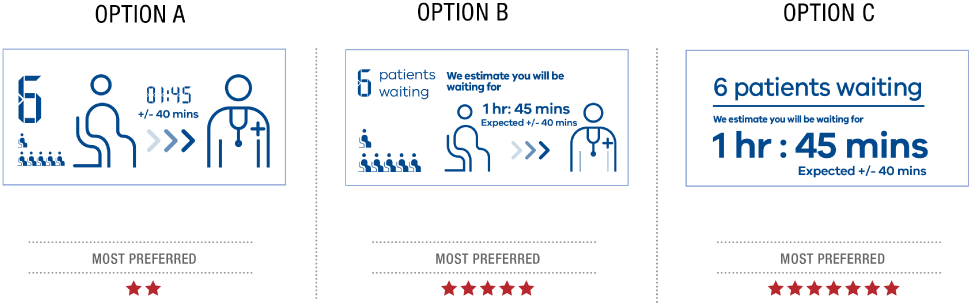

Most people felt that Option A was too obscure, although English speaking participants felt that it might be the easiest for those without English literacy skills to understand. Younger participants (teens to 50s) and people who had experience of English as a second language tended to favour the options that had graphics and symbols instead of text (Option B). English speaking participants *>* 70 years tended to prefer the text-only displays: “It’s more straightforward”, “It’s not so much information to take in. physical little pictures of patients; it’s really nothing there for me” (Option C).

### 4.1 How to convey ranges of estimates?

In Round 2 we retained the current time at the top to give context to the “Arriving now?” statement, and gave two options for presenting the time window to be seen.

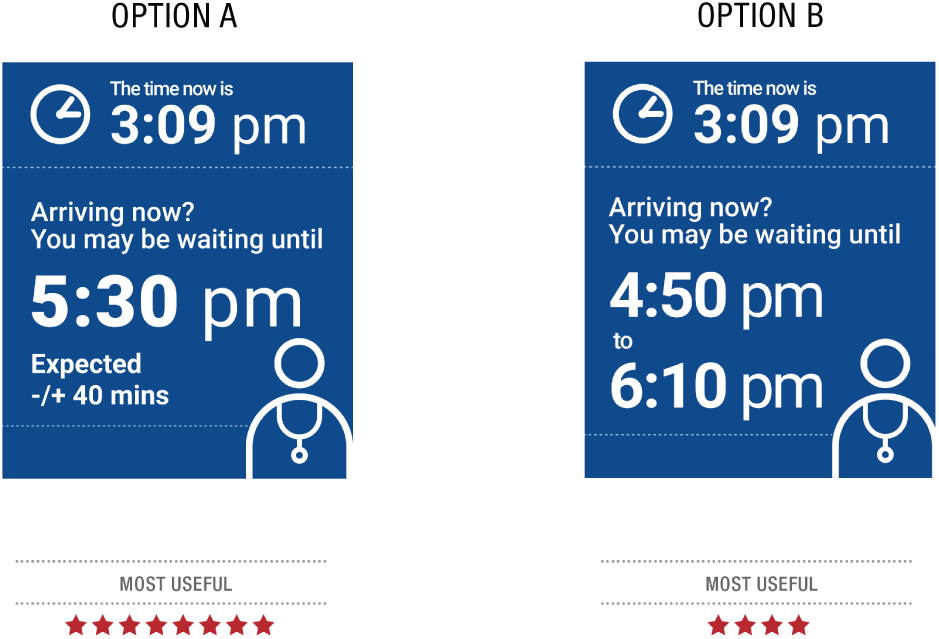

People also preferred the range to be presented with a single expected wait time and a +/-range, rather than having the range calculated for them (Option A). This was thought to be “more reassuring”, “less reading”, “less confusing”.

### 4.2 Qualitative or quantitative displays?

Taking inspiration from existing displays which show only a qualitative measure of “busyness” we next presented displays with varying levels of quantitative overlay:

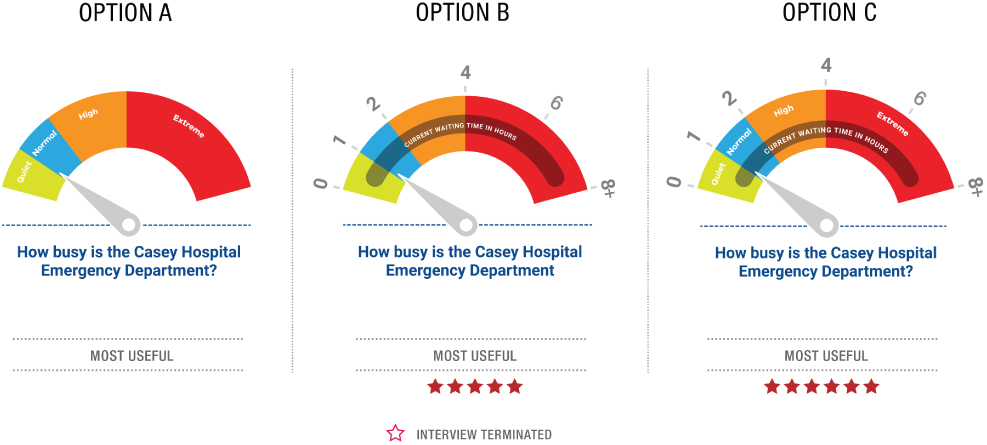

All participants preferred a ‘glancable’ pictogram display that also conveyed more detailed information about estimated wait times and the number of patients in the queue. Participants said that the barometer was most useful with both “current waiting time in hours” and “status” information on it (Option C). Less quantitative data made it less useful, so option A was the least preferred display.

### 4.3 Other patients waiting

From the previous set, we knew participants liked having information about the number of patients in the queue (numerical statement). Next we tested whether they would further appreciate a breakdown of the numbers of patients by urgency category. We displayed urgency patient icons as follows: Australasian Triage Scale (ATS) category 1 - critical/red, ATS 2 - high/orange, ATS 3 - medium/green and ATS 4 or 5 - blue/low icons in an Isotype display:

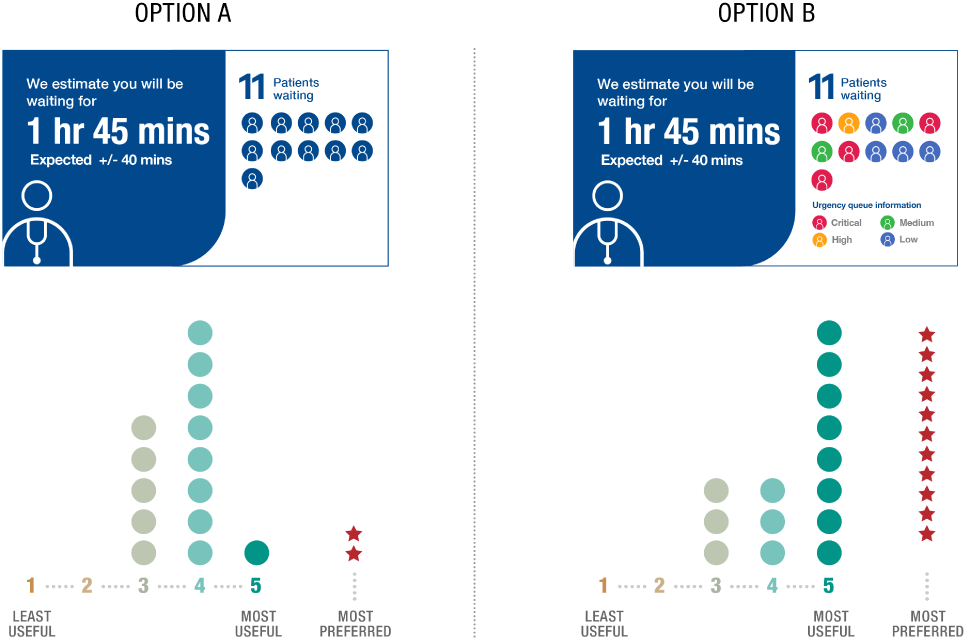

People felt that knowing others’ urgency would make them feel more reassured about their own waiting time and be able to place their situation in perspective. Participants all seemed to understand where their urgency would sit on this scale and be comfortable with this categorisation. In contrast, triage nurses and clerks raised concerns that some patients would dislike seeing that others might be classed as more urgent than themselves, potentially generating anger.

Thus, in Round 3, we tested two alternatives for the icon style:

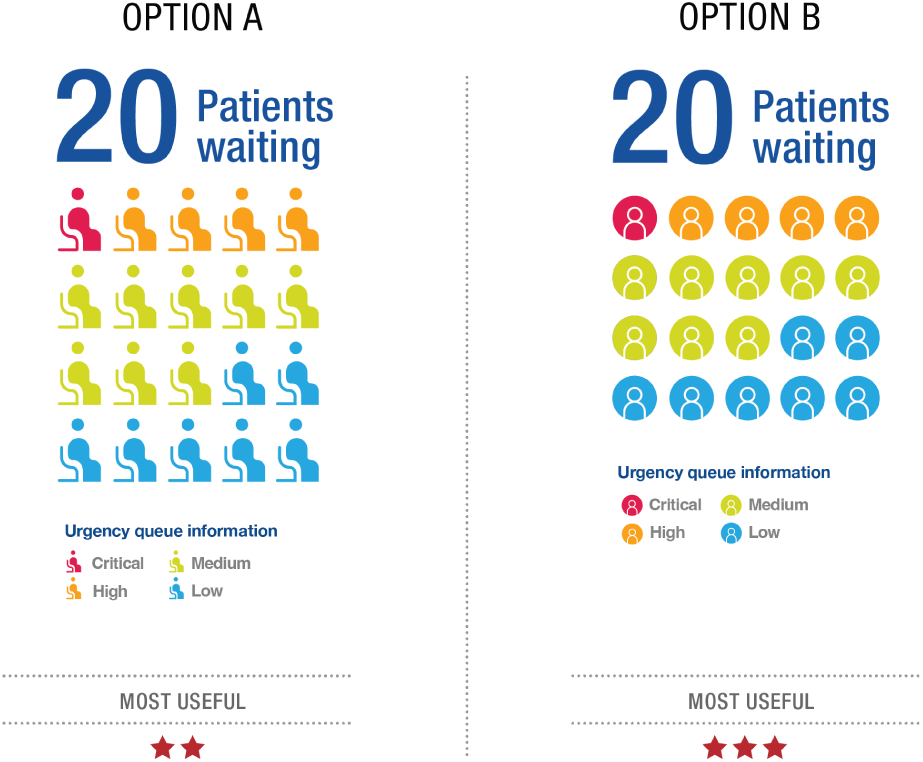

Initial preferences for which figure to use to display patients in queues was for the “round icons” (Option B) rather than the “seated icons” (Option A). Responses reflected personal design style preferences, rather than icon comprehension, and so either could be used and understood.

### 4.4 ED Capacity pictograms

Returning to the question of style of pictogram display of busyness, we tested three styles in Round 1:

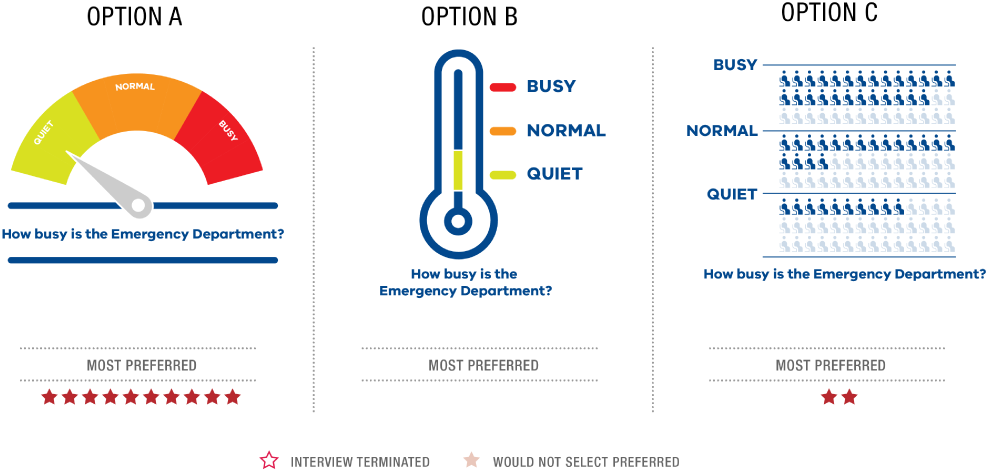

Participants recognised the “barometer” graphic (Option A) and they made a conceptual comparison to Australian “Fire Danger” signs. They regarded it as being clear, high impact and direct. Option A was the preferred option. People understood Option B, but felt that it lacked visual impact compared to the barometer. People were confused by Option C, for communicating ED busyness or capacity. They wondered if each person represented an exact number of patients or a quantity (e.g. 10 patients) waiting.

### 4.5 Combined element displays

Ultimately, the goal is to place all of these elements together in a single display. In our first attempt at a combined display (Round 1) we were concerned with the issue of how to support patients’ understanding of time, and we chose to test a standard data visualisation representation of events on a time line, showing current time, expected time to be seen and the most likely window around this expected time. With more space devoted to the timeline, we also had space to display additional arrival information. We felt that since this arrival information is one of the key inputs to the algorithm estimating wait time, it would also help to reassure patients trust in the estimate. The display exactly as shown to participants is in Fig. 1(right). Responses to the different elements of the display are shown in situ in the following infographic:

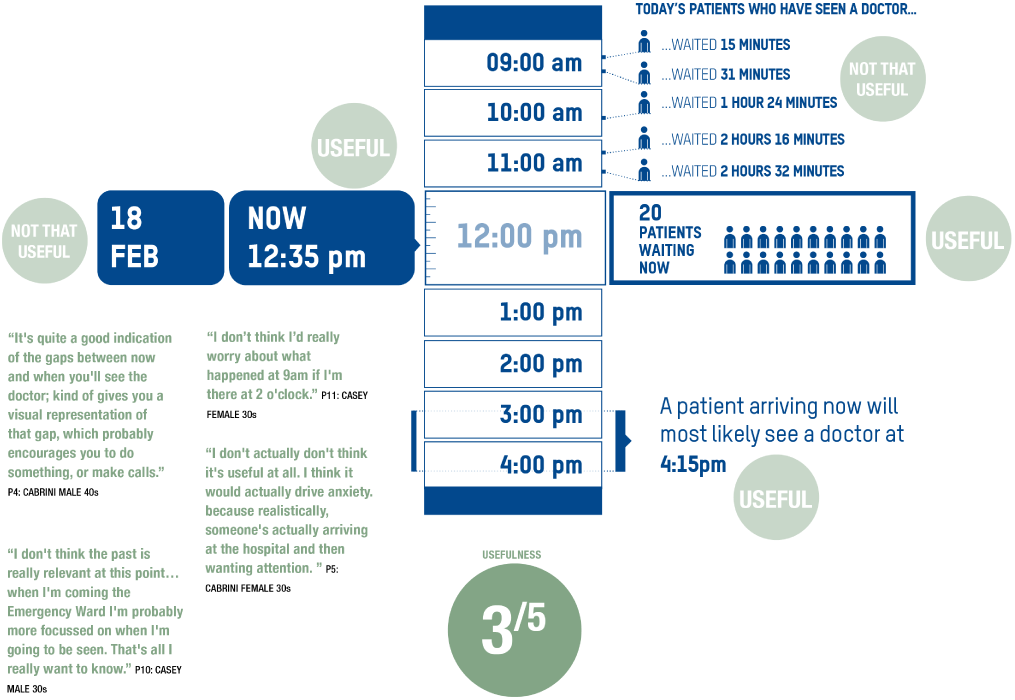

There were mixed responses to this style of display. People with higher levels of education and an interest in data-heavy displays liked this graphic. These people were interested in seeing the patterns of the day and how things “built up”. One third of participants felt it looked too busy. This may have been an effect of “visual load”, where efficient cognitive processing is hampered when displays are complex. Most preferred horizontal to vertical time progression displays. Participants in general found the lower half of the display useful, suggesting that the most useful parts are how many patients are waiting to be seen and the estimated waiting time for those that have just arrived at the ED: “I don’t think the past is really relevant at this point… when I’m coming the Emergency Ward I’m probably more focused on when I’m going to be seen. That’s all I really want to know.” (Participant 10). The upper half of the display made some people stressed and annoyed, with the realisation that if they had attended earlier, they would have had a shorter wait. The overall negative response led us to abandon the timeline display and the additional arrival information in subsequent “all-in-one” displays.

#### 4.5.1 Safety statements

The study team proposed multiple statements that could be placed on a display. Additional statements were added after discussion with triage nurses and clerks. A prioritisation process was undertaken amongst the research team, then statements were simplified. Participant understanding of the highest priority statements was tested (Fig. 7). The statements about queue management and safety were well understood by all participants. They did not suggest any wording changes. The highest priority statements were:

“Patients are seen in order of urgency, not arrival”

“If you feel worse, tell your triage nurse immediately”

“If you want to leave, tell us first”

#### 4.5.2 Order of images across a screen

Participants in Round 3 found the information presented with a blue background the most useful and wanted this to be seen first and on the left.(Fig. 6)

**Fig. 6.**
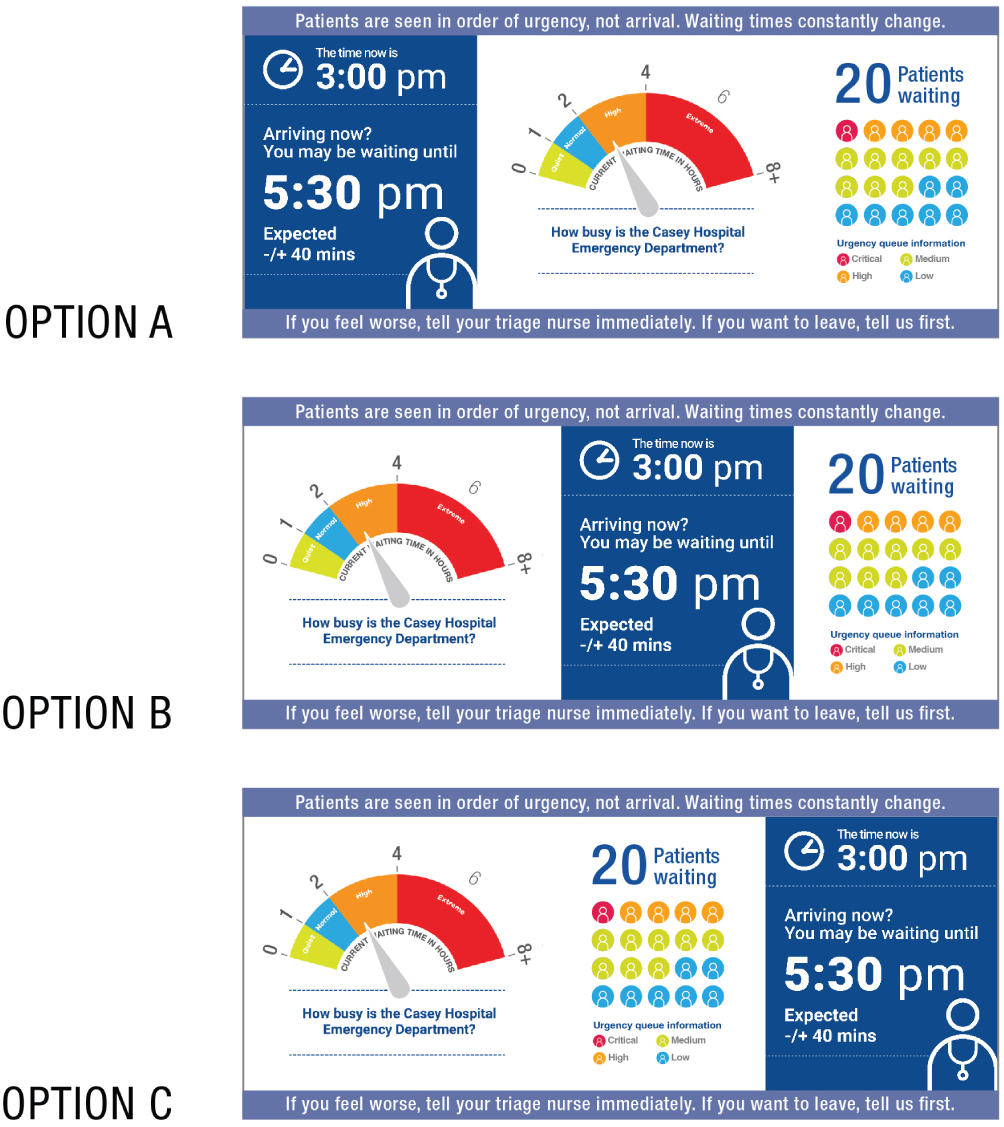
Left to right distribution of information

A more graphic, less word-dominant representation of the waiting process was recognised as being useful for people with less English comprehension, low literacy or cognitive impairment. Participants in these groups favoured options with infographic and pictographic information presentations, such as the “seated” patients waiting and visual depiction of “you may be waiting to see a doctor” (Fig. 7). Consequently, Fig. 7 was slightly more preferred than Fig. 8. With graphic elements rather than textual aspects dominating in this version, participants felt they could access the information that was most important to them – the overall ED busyness and an indication of the number of people waiting to be seen by a doctor.

**Fig. 7.**
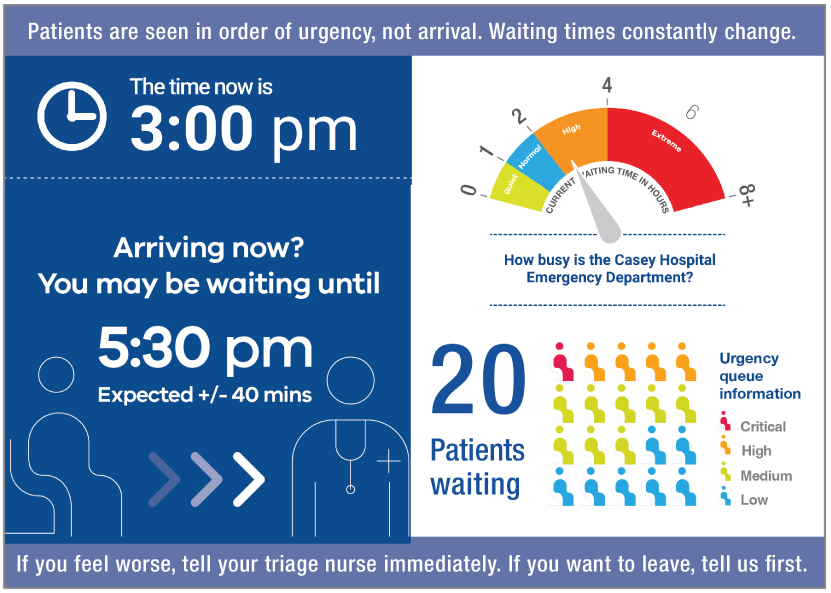
More images and less words preferred by CALD participants and those with lower English literacy.

**Fig. 8.**
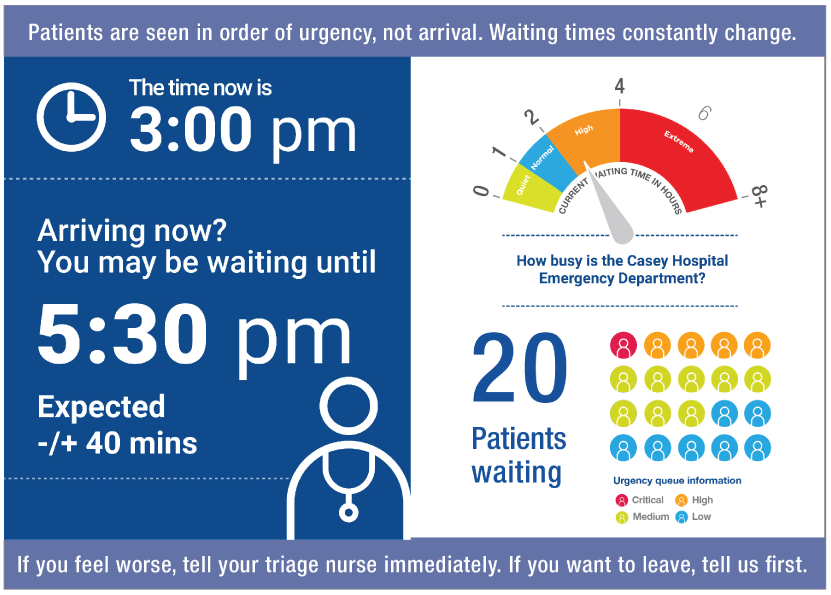
More words and less images were preferred by those with higher educational attainments

#### 4.5.3 Two or Three columns available

From the above feedback, it was determined that current time and expected time to be seen are essential information, as well as a general statement of busyness summarised in the barometer style. Participants preferred to retain urgency queue information. At the time of writing it was unclear the dimensions that would be available on the physical waiting room displays as well as the website deployment. We therefore have recommendations for both two and three column versions.

Our final (Round 4) comparative test for the two column version revisited stylistic elements used in the previous round, simpler or more figurative graphic elements:

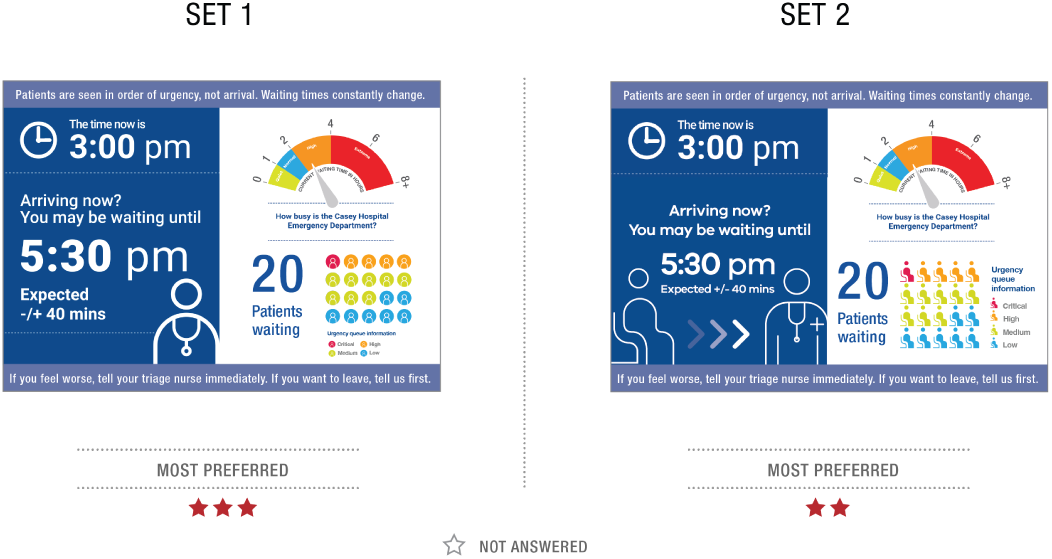

If two columns of space were available, they would prefer Set 1 from the above for their display, with the blue section on the left. They were happy to accept Set 2 as an alternative.

Overall preference, however, was for a three column display. This makes sense, since our final design features three basic elements (time statement, busyness barometer and urgency queue info) a display separating each of these into their own column is recommended if there is enough horizontal space. If three columns are available on a screen, then the preferred display is shown in Fig. 9.

**Fig. 9.**
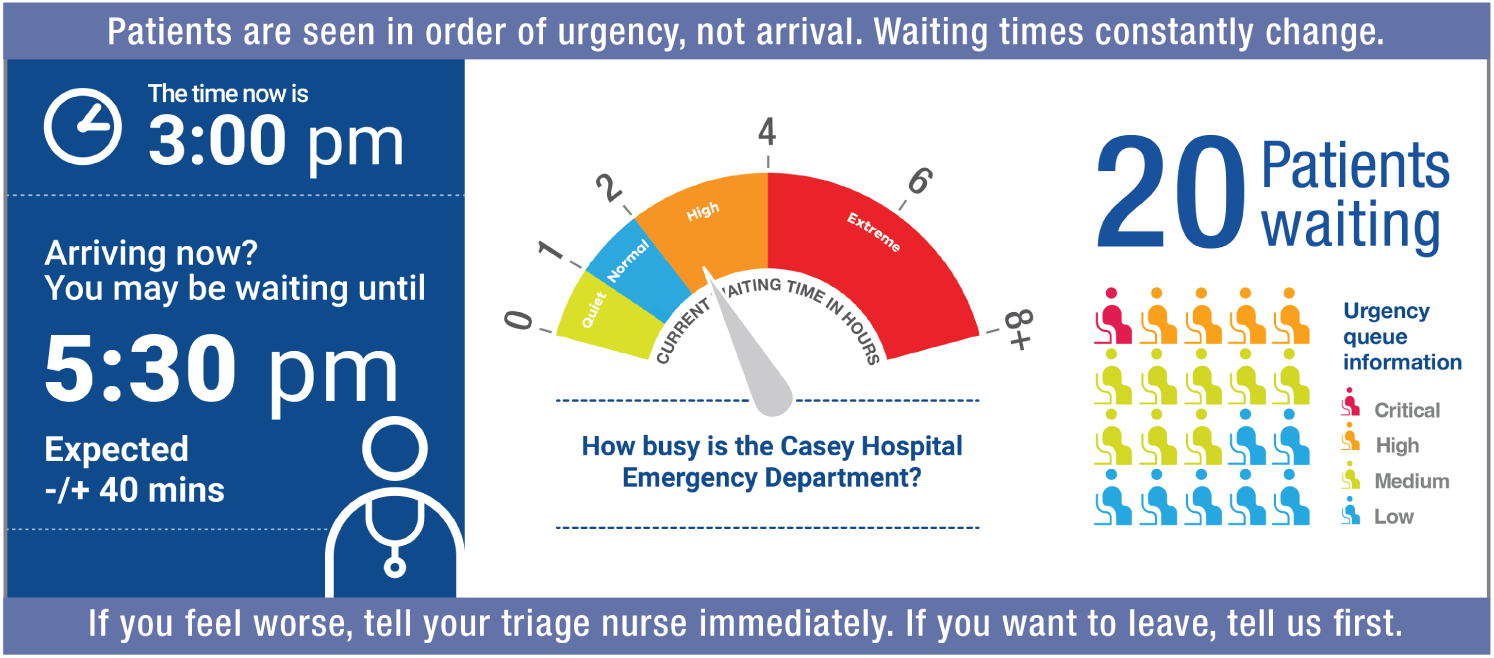
Best version of 3-column display

## 5 Discussion

This study used RITE user experience methodology to co-design emergency wait time prediction displays for patients and families, using screens in ED waiting rooms. RITE methodology worked well for us as a pragmatic approach, making the most of our limited access to real patients in ED waiting rooms during a time of COVID causing hospital access issues as well as stress on both patients and staff. Rather than traditional controlled studies carefully isolating particular factors for statistical analysis across task-based measures, we tested multiple alternative design elements in each round. To tease apart the most successful aspects of each design alternative, we relied on extensive qualitative discussion and feedback. Elements deemed successful were carried through to subsequent rounds, and ultimately we were able to converge on a design recommendation with detailed justification for each of its components, as follows.

Participants wanted to see the following: current time, (both analogue/numbers and clock/figure); expected wait time data on the left; a calculated time to be seen and a range (Expected +/-xx minutes/hours). The calculated time to be seen within an expected +/-range was regarded as the most useful information. They also wanted a barometer “busyness” display containing quantitative data; a quantitative statement of how many patients are already queuing and an image showing queue urgency, with icons for patients waiting and colours for urgency. Participants understood the safety and queue management statements. Complex, multi-leveled visualisations were sought by some and rejected by others.

At the conclusion of our iterative design process, Fig. 9 is our recommendation, and the version that will hopefully be deployed in 2022 in Victorian ED wait rooms and websites. It conveys the most straightforward statement about current and expected time to be seen, rather than wait duration as per Sect. 4.0.1 or a range of times as per Sect. 4.1. The graphical elements are placed to the right of the text statement of time to be seen (as per Sect. 4.5.2). These graphical elements seek to augment rather than replace the text. The barometer pictogram (centre) is adapted from existing displays used in one Victorian ED wait room (e.g. Fig. 2) but it is based on more meaningful information, i.e. an estimate of wait time in hours, rather than just an abstract “degree of business”. To the right we include the Isotype display of urgency queue break down. While hospital administrators were concerned about giving this level of information to patients, the overwhelming feedback from patients is that it reassured them that resources were being allocated where most needed, as determined in Sect. 4.3.

## 6 Conclusion, Recommendations and Future Work

This may be the first healthcare study to investigate temporal uncertainty displays for consumption by the general public. Design and visualisation studies intended for healthcare applications tend to focus on discrete aspects of perceptually effective visual displays for clinicians’ use [26], however studies of public-facing, real-time waiting information in healthcare settings—particularly those with a user-centred focus—are currently lacking. Chung et al. describe previous methods of conveying uncertainty visually, including modifying or animating graphical attributes, adding artefacts to displays, using non-visual techniques (e.g. sound) or allowing user interaction with data features (such as a mouse hovering over a data-point, triggering a pop-up explanation) [6]. Despite this variety of possible methods, studies of the impact of these visualisations on end-users and hence their influence on decision making are limited. Grewal et al. report about 50 visualisation applications available to convey temporal uncertainty in their UncertaintyViz Browser, none related to health [24]. Tominski et al identify five publications where time is visualized in health, none of these visualisations are for patient consumption [15]. This is remarkable given that 4-8 percent of national Gross Domestic Product is spent on healthcare [43] and wait durations are highly politicised. Most people in most places wait for consultations and treatment. Treatment settings where wait time visualisations would be helpful to patients, include acute (e.g. emergency), chronic (e.g. specialist clinics), and elective care (e.g. surgery). In contrast, other general public domains, such as transport, have published a handful of manuscripts [25, 44].

We recommend that if an entire region is starting to display wait times, the displays are all similar. In particular, the scales (such as on the graphic displaying hospital ED “busyness”) should all be the same to allow patients to compare one ED to another. Eventually, as information technology capabilities improve in health systems, individualised displays can be offered, perhaps to a personal mobile phone, where the user could vary accessibility features such as audio data delivery, language, font size, and color schemes to meet their needs.

The introduction of health technology and the visualisation of data has the potential to exacerbate existing healthcare access inequities, worsening what has been called “Techquity” [7, 35]. In the last two years, COVID-19 has caused transformative and disruptive change in healthcare delivery, with a rapid increase in the use of technology, and remote telehealth consultations at the expense of face-to-face attendances. This has reduced the ability of under-served groups to access healthcare and has widened inequality gaps [10]. Compounding this disadvantage, when AI is introduced, unfair prediction models are often built and deployed, using retrospective data from biased systems. This has been demonstrated in a wide range of domains such as health [14], finance [30], and corrections [12]. The models deprioritize less favoured groups such as women, ethnic minorities or older people. This can then exacerbate bias with unfair outputs [16,20]. In the context of wait time predictions and emergency care, the interpretation and use of wait time visualisations is particularly important for First Nations people, such as Australia’s Aboriginal and Torres Strait Islanders, who have double the rate of leaving EDs without being seen [3, 17, 38]. Further research using RITE methodology to design visual communications should prioritise the needs of under-served populations with high rates of leaving EDs without being seen, who may have the most to gain from having visualisations meet their needs.

This study was conducted in a city in a high-income, largely English-speaking country with participants with varied data literacy skills. It is likely that displays for populations in countries where data is usually displayed as symbols might prefer more icons or images and fewer words. Preferences for some of the images will be related to previous familiarity for graphical information displays – such as the fire danger barometer in Australia. Testing images on site in an ED with small numbers of participants (n=36) means that some patients, for example those with behavioural emergencies and families of patients who are critically unwell have not yet provided feedback. The impact of these displays on patients when wait times exceed predicted times has not been determined by this study.

In conclusion, this study demonstrates that valuable consumer input can be rapidly obtained and deployed into design iterations to visualise health data displays that align with end user needs and expectations.

## Data Availability

Data are not available due to privacy and consent concerns.

